# Loss of Factor H family proteins associates with meningococcal disease severity

**DOI:** 10.1101/2021.02.05.21251142

**Authors:** Anna E. van Beek, Richard B. Pouw, Victoria J. Wright, Neneh Sallah, David Inwald, Clive Hoggart, Mieke C. Brouwer, Rachel Galassini, John Thomas, Leo Calvo-Bado, Colin Fink, Ilse Jongerius, Martin Hibberd, Diana Wouters, Michael Levin, Taco W. Kuijpers, on behalf of the EUCLIDS consortium.

## Abstract

*Neisseria meningitidis*, the causative agent of meningococcal disease (MD), evades complement-mediated clearance upon infection by ‘hijacking’ the human complement regulator factor H (FH). The FH protein family also comprises the homologous FH-related (FHR) proteins, hypothesized to act as antagonists of FH, and FHR-3 has recently been implicated to play a major role in MD susceptibility. Here, we show that, next to FH and FHR-3, the circulating levels of all FH family proteins are equally decreased during pediatric MD. We did not observe a specific consumption of FH or FHR-3 by *N. meningitidis* during the first days of infection and the levels recovered over time. MD severity associated predominantly with a loss of FH. Strikingly, loss of FH and FHRs associated strongly with renal failure, suggesting insufficient protection of host tissue by the FH protein family. Retaining higher levels of FH family proteins may thus limit tissue injury during MD.

## INTRODUCTION

*Neisseria meningitidis* is a Gram-negative commensal bacterium carried in the nasopharynx by up to 24% of the population [1]. Upon infection, it can cause meningococcal meningitis and/or septicemia, collectively called meningococcal disease (MD). MD is a severe, debilitating and life-threatening disease with an occurrence of 2 – 20 per 100,000 in developed countries [2,3]. The complement system plays a major role in preventing MD, exemplified by deficiencies in components of its terminal pathway that are associated with recurrent *N. meningitidis* infection [4,5].

*N. meningitidis* is known to exploit human complement regulator factor H (FH) to avoid complement-mediated clearance [6,7]. FH is a glycoprotein circulating in plasma at around 300 µg/mL [8]. It is the major regulator of the alternative pathway and is composed of 20 complement control protein (CCP) domains. FH is crucial in protecting human cells from complement-mediated damage and contains two regions involved in the binding to human cells, located in CCP6 to CCP8 and in CCP19 and CCP20 [9]. *N. meningitidis* expresses various proteins to recruit FH to its surface [6,7,10–12]. FH-binding protein (fHbp) plays a dominant role in evading complement-mediated clearance upon infection [13]. This lipoprotein binds FH at CCP6 and CCP7, while leaving the complement regulatory capabilities of FH intact [14]. The ‘hijacking’ of FH aids *N. meningitidis* in avoiding complement-mediated clearance, prolonging its survival in human circulation [6,7,15].

The FH protein family is encoded in tandem in the *CFH-CFHR* locus. *CFH* encodes FH and its short splice variant, FH like-1 (FHL-1), while the five *CFHR* genes encode the homologs FH-related (FHR)-1, FHR-2, FHR-3, FHR-4A and FHR-5 [16]. FHR-1, FHR-2 and FHR-5 circulate in blood as dimers, with FHR-1 and FHR-2 also forming heterodimers [17,18]. The FHRs have high sequence identity to the ligand binding regions of FH (CCP6-8 and CCP19-20), but lack CCP domains homologous to FH CCP1-4, which are involved in complement regulation [19,20]. Therefore, FHRs are hypothesized to compete with FH binding to cellular surfaces, enhancing complement activation [16].

A previous genome-wide association study identified that, apart from SNPs in *CFH*, gene variations in *CFHR3* were also found to associate with MD susceptibility [21]. This was the first indication that the FHRs might play a role in MD, with FHR-3 as the most promising candidate to compete with FH. FHR-3 has the highest sequence identity with FH CCP6 and CCP7 (91% and 85%, respectively), and was found to bind to fHbp in vitro, competing with FH for binding [22].

Since we recently developed FHR-specific ELISAs, we were now able to make the translation from genetics and in vitro data towards the study of FH family proteins and their levels during the acute stage of MD. We previously reported that the levels of FHRs are 10 – 100 fold lower in comparison to FH during steady-state [8,18,23]. However, the levels of FH and the FHRs may very well be altered during an episode of acute MD, possibly affecting their ratio and changing the balance of alternative pathway activation and regulation.

In this study, we analyzed the serum levels of FH and all FHRs from a cohort of pediatric MD patients during the acute stage of disease in relation to *N. meningitidis* serogroup, diagnosis and severity parameters and compared these with levels during convalescence in surviving patients. We report here that not only FH and FHR-3, but plasma concentrations of all FH family proteins are greatly decreased during the acute phase of MD. However, predominantly low FH plasma concentrations are associated with the severity of MD.

## RESULTS

### Study cohort

Serum samples were available from 106 children with MD (**Table 1**). The patient age ranged from 0.1 – 16 years at admission with a median age of 2.9 years with equal gender distribution. Convalescent samples were drawn 10 – 2011 days after infection, with a median of 65 days. Acute stage samples were obtained at the first or second day of hospitalization. In seventeen patients, samples were also obtained during subsequent days. Fourteen patients (13%) were diagnosed with localized meningococcal meningitis, 75 (71%) with meningococcal septicemia, and seventeen (16%) with both (i.e. proven septicemia and meningitis). Fifteen out of the 106 patients did not survive. Death occurred at a median of one day (range 0 – 11). *N. meningitidis* serogroup was successfully typed in 69 cases, with serogroup B being the most prevalent (43 cases, 61%), followed by serogroup C (24 cases, 34%) and single cases of serogroup A and W135.

**Table 1.**
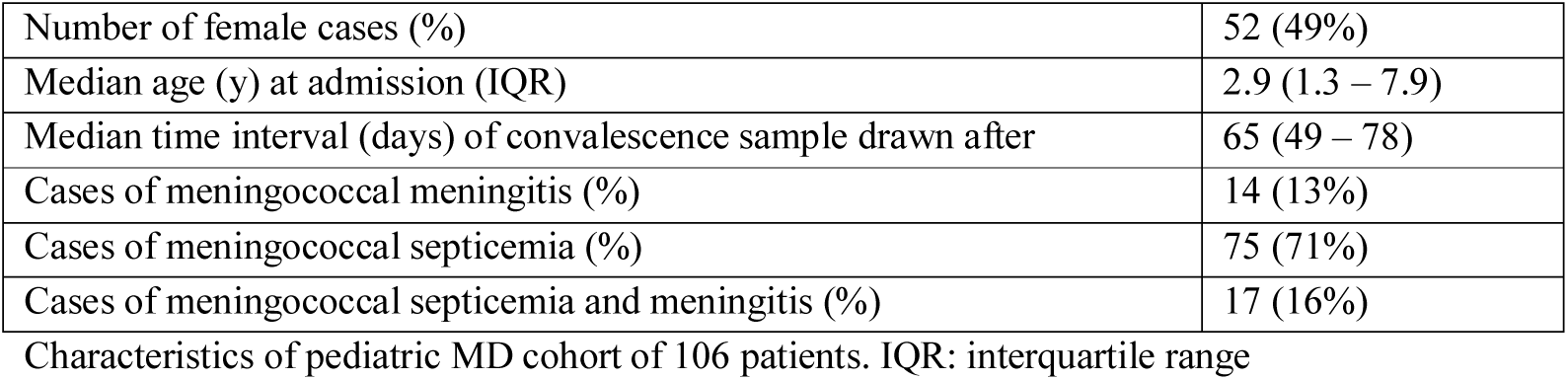
Patient characteristics.

### FH and FHR protein levels decrease equally during MD

We assessed the levels of FH and all FHRs in the first sample drawn at the acute stage and at convalescence from MD patients, using in-house developed ELISAs (**Figure 1a and 1b**) [8,18,23]. FH, FHR-1/1, FHR-1/2, FHR-2/2 and FHR-5 levels at convalescence were comparable to those of healthy children (**Table 2**) [24]. Levels of FHR-3 and FHR-4A were found to be higher at convalescence than FHR-3 and FHR-4A levels in healthy children. Two children appeared to carry a homozygous *CFHR3/CFHR1* deletion, as evidenced by the lack of either protein in their convalescent sample. No apparent *CFHR3/CFHR1* deletion was found among the non-survivors, based on their acute stage FHR-3 and FHR-1/1 levels.

**Table 2.**
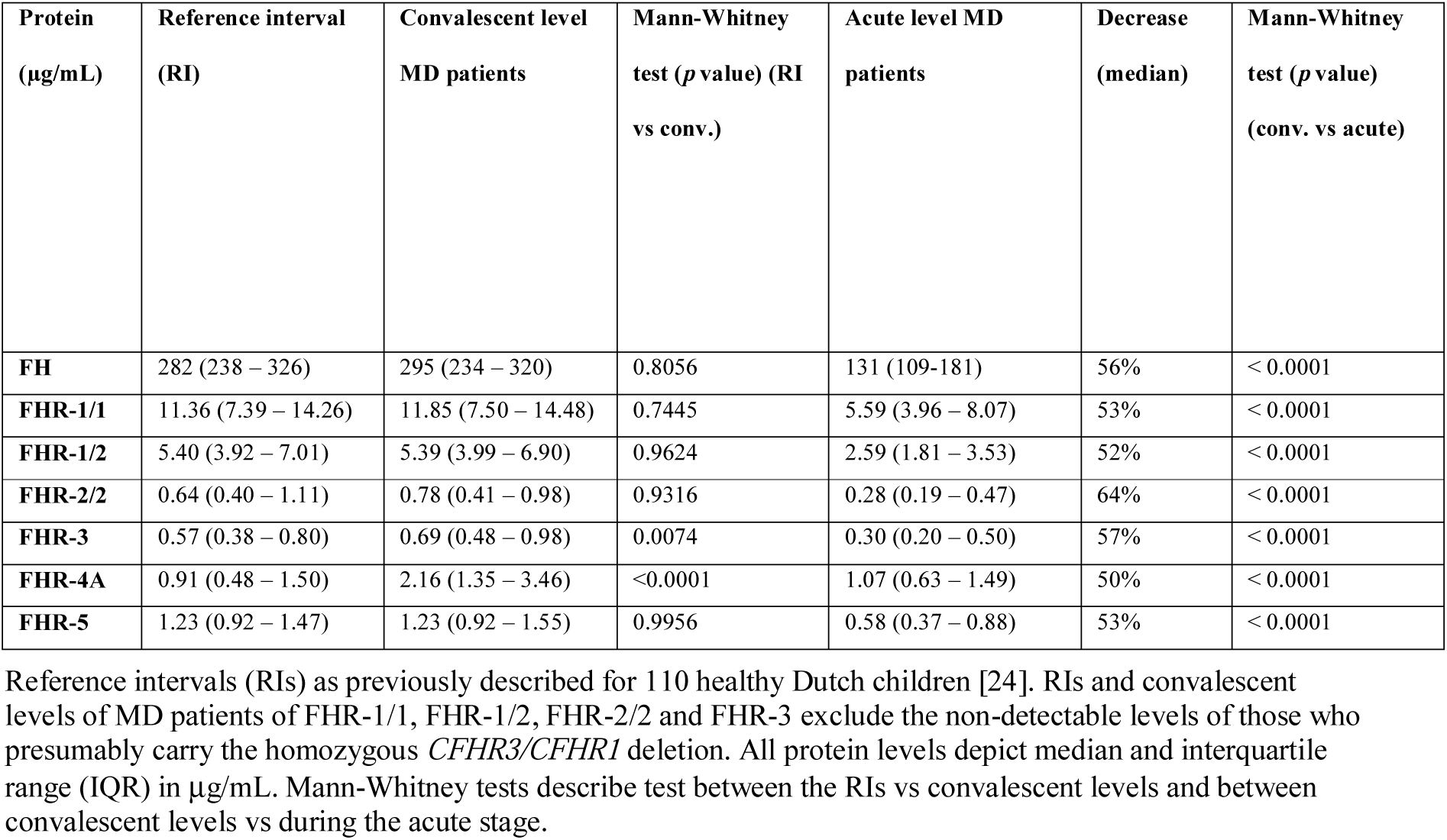
FH protein family ranges during MD.

| Protein<br>(µg/mL) | Reference interval<br>(RI) | Convalescent level<br>MD patients | Mann-Whitney<br>test (*p* value) (RI<br>vs conv.) | Acute level MD<br>patients | Decrease<br>(median) | Mann-Whitney<br>test (*p* value)<br>(conv. vs acute) |
| --- | --- | --- | --- | --- | --- | --- |
| **FH** | 282 (238 – 326) | 295 (234 – 320) | 0.8056 | 131 (109-181) | 56% | < 0.0001 |
| **FHR-1/1** | 11.36 (7.39 – 14.26) | 11.85 (7.50 – 14.48) | 0.7445 | 5.59 (3.96 – 8.07) | 53% | < 0.0001 |
| **FHR-1/2** | 5.40 (3.92 – 7.01) | 5.39 (3.99 – 6.90) | 0.9624 | 2.59 (1.81 – 3.53) | 52% | < 0.0001 |
| **FHR-2/2** | 0.64 (0.40 – 1.11) | 0.78 (0.41 – 0.98) | 0.9316 | 0.28 (0.19 – 0.47) | 64% | < 0.0001 |
| **FHR-3** | 0.57 (0.38 – 0.80) | 0.69 (0.48 – 0.98) | 0.0074 | 0.30 (0.20 – 0.50) | 57% | < 0.0001 |
| **FHR-4A** | 0.91 (0.48 – 1.50) | 2.16 (1.35 – 3.46) | <0.0001 | 1.07 (0.63 – 1.49) | 50% | < 0.0001 |
| **FHR-5** | 1.23 (0.92 – 1.47) | 1.23 (0.92 – 1.55) | 0.9956 | 0.58 (0.37 – 0.88) | 53% | < 0.0001 |

Reference intervals (RIs) as previously described for 110 healthy Dutch children [24]. RIs and convalescent levels of MD patients of FHR-1/1, FHR-1/2, FHR-2/2 and FHR-3 exclude the non-detectable levels of those who presumably carry the homozygous *CFHR3/CFHR1* deletion. All protein levels depict median and interquartile range (IQR) in µg/mL. Mann-Whitney tests describe test between the RIs vs convalescent levels and between convalescent levels vs during the acute stage.

**Figure 1.**
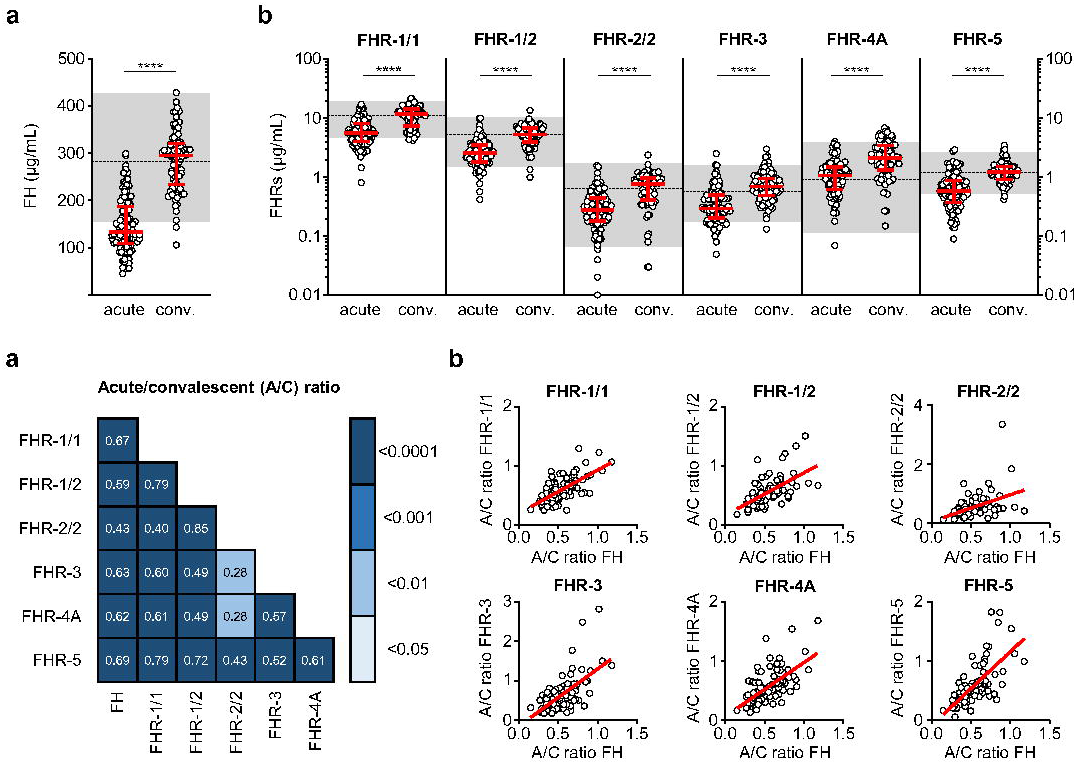
FH family protein levels are low at the acute stage of MD. **(a**,**b)** Differences in FH, FHR-1/1 homodimers, FHR-1/2 heterodimers, FHR-2/2 homodimers, FHR-3, FHR-4A and FHR-5 homodimers as assessed at the acute stage (samples obtained during the first or second day of hospitalization, *n* = 106) compared to levels at convalescence (*n* = 91). Two children appeared to carry a homozygous *CFHR3/CFHR1* deletion, as evidenced by the lack of either protein in their convalescent sample. They were excluded from the analysis for FHR-1/1, FHR-1/2, FHR-2/2 and FHR-3 (*n* = 104 and *n* = 89 for acute stage and convalescence). Acute serum samples comprised 88 samples drawn at day 1 and 18 samples drawn at day 2 of hospitalization, for patients of whom no day 1 sample was available. Levels of FHR-2/2 were calculated based on FHR-1/1 and FHR-1/2 levels. Shaded area indicates 95% range in healthy patients, with dashed line indicating the median. Scatter dot plots depict median and interquartile range (IQR) as red lines. Statistical significance was tested using a Mann-Whitney test. **(c)** Correlations (*r*) between the relative decreases of FH family proteins (ratios between acute and convalescent levels, by dividing acute levels over convalescent levels) were assessed using Pearson’s measure of association, followed by the Benjamini-Hochberg procedure to control for the false discovery rate (FDR, set to 0.05). Blue shades indicate, from light to dark: *p* < 0.05; *p* < 0.01; *p* < 0.001; and *p* < 0.0001. (**d**) Examples of correlations in (**c**), showing relative decrease in FH levels versus relative decrease in FHR levels. A:C ratio: acute:convalescent ratio; Conv.: convalescent.

At the acute stage, both FH and all FHRs were markedly decreased, with median levels 50-64% lower than found at convalescence (**Figure 1, Table 2**). In contrast to all other FH family proteins, the acute stage FHR-4A levels, while being significantly decreased compared to those found at convalescence, were not significantly lower compared to the normal range found in healthy children. The relative decreases of all FH family proteins strongly correlated with each other, indicating a similar underlying mechanism (**Figure 1c and 1d**). Of note, the FH and FHR ELISAs were not affected by the possible presence of circulating fHbp in the acute samples (**Supplementary Figure 1**).

### FH family proteins show different kinetics following the acute stage of infection

After having established that all FH family proteins were decreased proportionally during the acute phase of infection, we investigated the changes in concentration on subsequent days in a subset of seventeen patients. In contrast to the equal decrease upon infection, recovery of the FH family proteins in the subsequent days differed from each other (**Figure 2a and 2b**). The median FH levels remained low during the first days of hospitalization, but showed a sign of recovery to normal levels at day four. In contrast, levels of FHR-1/1, FHR-1/2, FHR-2/2 and FHR-3 showed a quicker recovery. FHR-4A levels did not change during the first four days of hospitalization, remaining at approximately 50% of convalescent levels. Blood levels of FHR-5 recovered more steadily, similar to FH. The decrease in serum levels at the acute stage of MD was not unique to the FH family proteins, since total IgG levels, which were measured as a reference protein of similar molecular weight as FH, showed a similar decrease and dynamics (**Figure 2c**) [25].

**Figure 2.**
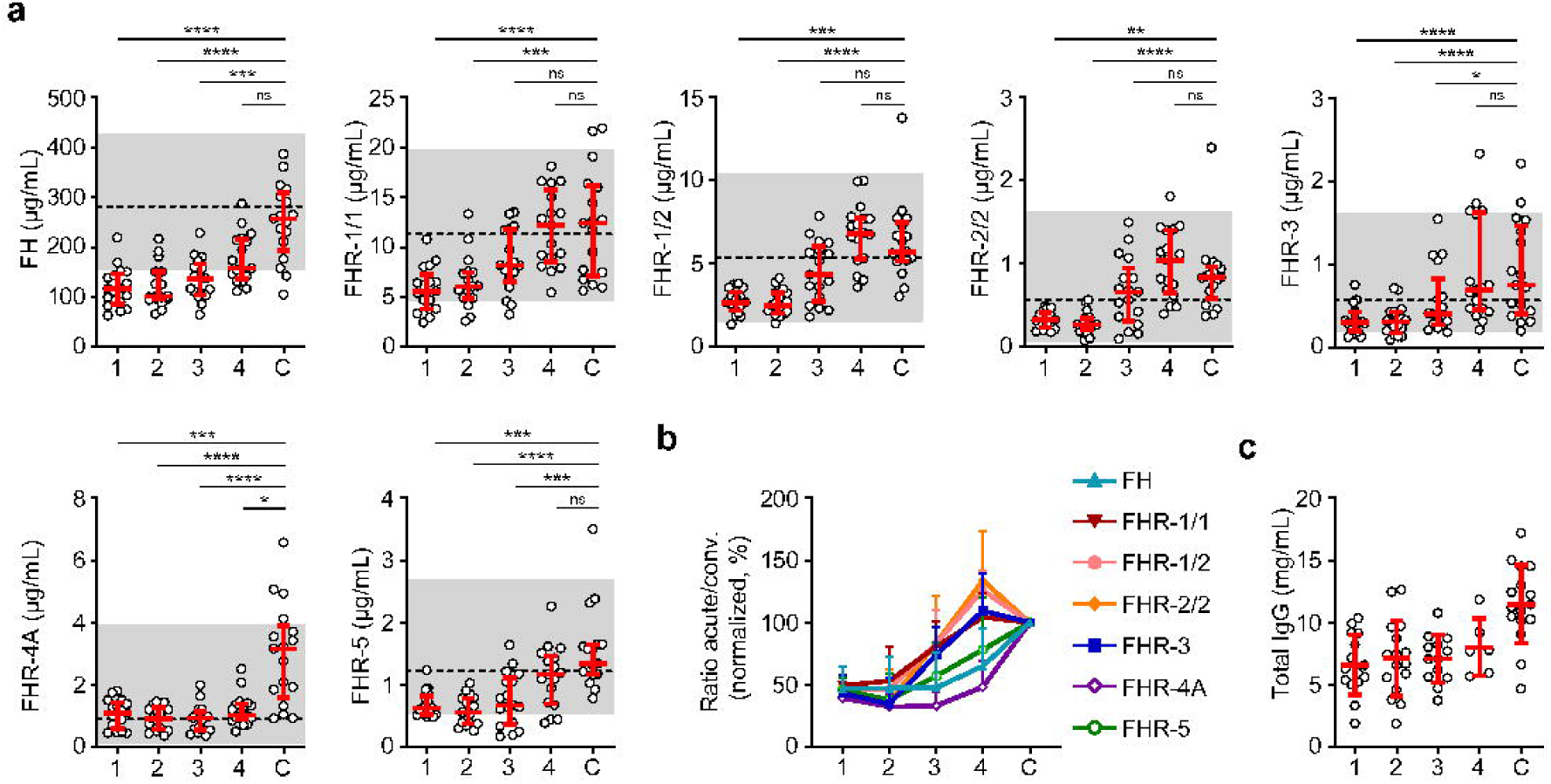
FH family protein level dynamics during the acute stage of MD. **(a)** FH family protein levels in paired samples (*n* = 17) as assessed during the first four days of infection (day 1 until day 4), compared with the concentration at convalescence (C). Shaded area indicates 95% range in healthy patients, with dashed line indicating the median. Friedman test followed by Dunn’s multiple comparisons test, with every acute stage dataset compared to the levels found at convalescence. ****: *p* < 0.0001; ***: *p* < 0.001; **: *p* < 0.01; *: *p* < 0.05; ns = not significant. **(b)** FH family protein levels as in **(a)**, normalized to the levels found at convalescence. **(c)** Total IgG levels in unpaired samples (maximum *n* = 16) as assessed during the first four days of infection and at convalescence. The 95% range in healthy patients is not depicted, due to variability of total IgG levels during childhood. Lines depict median and IQR.

### FH and FHR levels associate with clinical and laboratory parameters

Next, we analyzed whether the low FH and FHR levels at the acute stage were related to the classification or severity of MD. Levels of FH, FHR-1/1, FHR-1/2, FHR-3 and FHR-4A were lower in patients diagnosed with septicemia in comparison to those with meningitis alone (**Figure 3**). Levels of FHR-2/2 were comparable between the patient groups, while FHR-5 levels were lower in patients who were diagnosed with septicemia alone when compared to meningococcal meningitis with accompanying septicemia (**Figure 3**).

**Figure 3.**
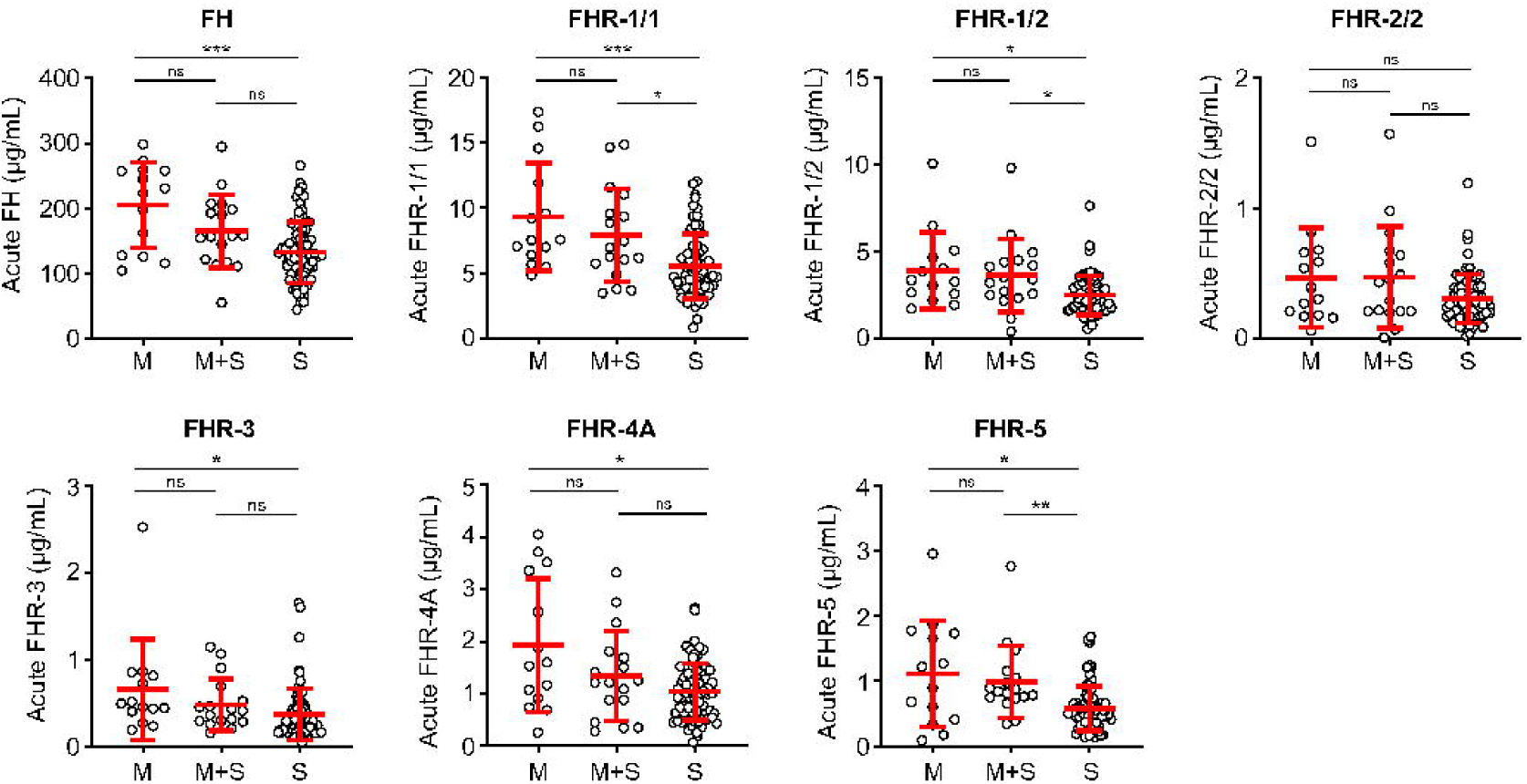
FH family proteins per clinical syndrome. Protein levels of FH, FHR-1/1, FHR-1/2, FHR-3, FHR-4A and FHR-5 at the acute stage (first sample obtained during hospitalization, *n* = 106), according to the diagnosed clinical syndrome: meningococcal meningitis (M, *n* = 14), meningococcal septicemia (S, *n* = 75), or both (M+S, *n* = 17). Statistical significance was tested using a Kruskal-Wallis test, followed by a Dunn’s multiple comparisons test. Lines depict median and IQR. **: *p* < 0.01; *: *p* < 0.05; ns = not significant.

We also assessed whether the FH and FHR levels at acute stage correlated with the clinical parameters WCC, platelet count, aPTT, INR, fibrinogen, CRP, potassium, base excess, lactate, the severity scores PIM and GMSPS, and the bacterial load (**Table 3, Figure 4**). Overall, all correlations indicated a negative effect of low acute stage FH and/or FHR levels, i.e. low protein levels correlating with more adverse clinical values. Acute stage FH levels correlated with most clinical parameters, with only potassium, lactate and the PIM score showing no significant correlation with FH. Of the FHRs, FHR-1 and FHR-5 correlated most with various clinical parameters. There was a striking correlation between low levels of all FH family proteins and base excess. Although reduced base excess correlated with increased lactate levels in the patients (*r* = −0.56, *p* < 0.0001) the FH family protein levels were not correlated with lactate. Although both base excess and lactate are markers of impaired perfusion and shock, the correlation with base excess and not with lactate may have been influenced by the increase in base excess associated with resuscitation with high chloride-containing fluids (normal saline or 5% albumin), which cause a worsening of base excess due to hyperchloremic acidosis [26]. FH family protein levels were lowest in those patients receiving renal support (**Figure 5, Supplementary Figure 2**).

**Table 3.** Clinical and laboratory parameters.

| Parameter | Normal |  | MD patients |
| --- | --- | --- | --- |
|  |  | *n* | Median (IQR) |
| WCC (*10<sup>9</sup>/L) | 4.0 – 11.0 | 105 | 9.3 (3.7 – 23.5) |
| Platelet count | 150 – 400 | 105 | 196 (132 – 256) |
| aPTT (s) | 30 – 40 | 99 | 45.9 (36 – 61.9) |
| INR | 0.8 – 1.2 | 94 | 1.6 (1.3 – 1.8) |
| Fibrinogen (g/L) | 1.5 – 4.0 | 84 | 3.1 (1.9 – 4.5) |
| CRP (mg/L) | 5 – 10 | 93 | 92 (54 – 157) |
| Potassium (mM/L) | 3.5 – 5.0 | 103 | 3.5 (3.2 – 3.9) |
| Base Excess (mEq/L) | -2 – 2 | 104 | -7 (-10 – -5) |
| Lactate (mmol/L) | 0.5 – 1 | 56 | 1.6 (0.9 – 3.7) |
| PIM | 0 | 100 | 4.9 (2.4 – 14.0) |
| GMSPS | 0 | 106 | 10 (7 – 12) |

*n*: number of informative cases; IQR: interquartile range.

**Figure 4.**
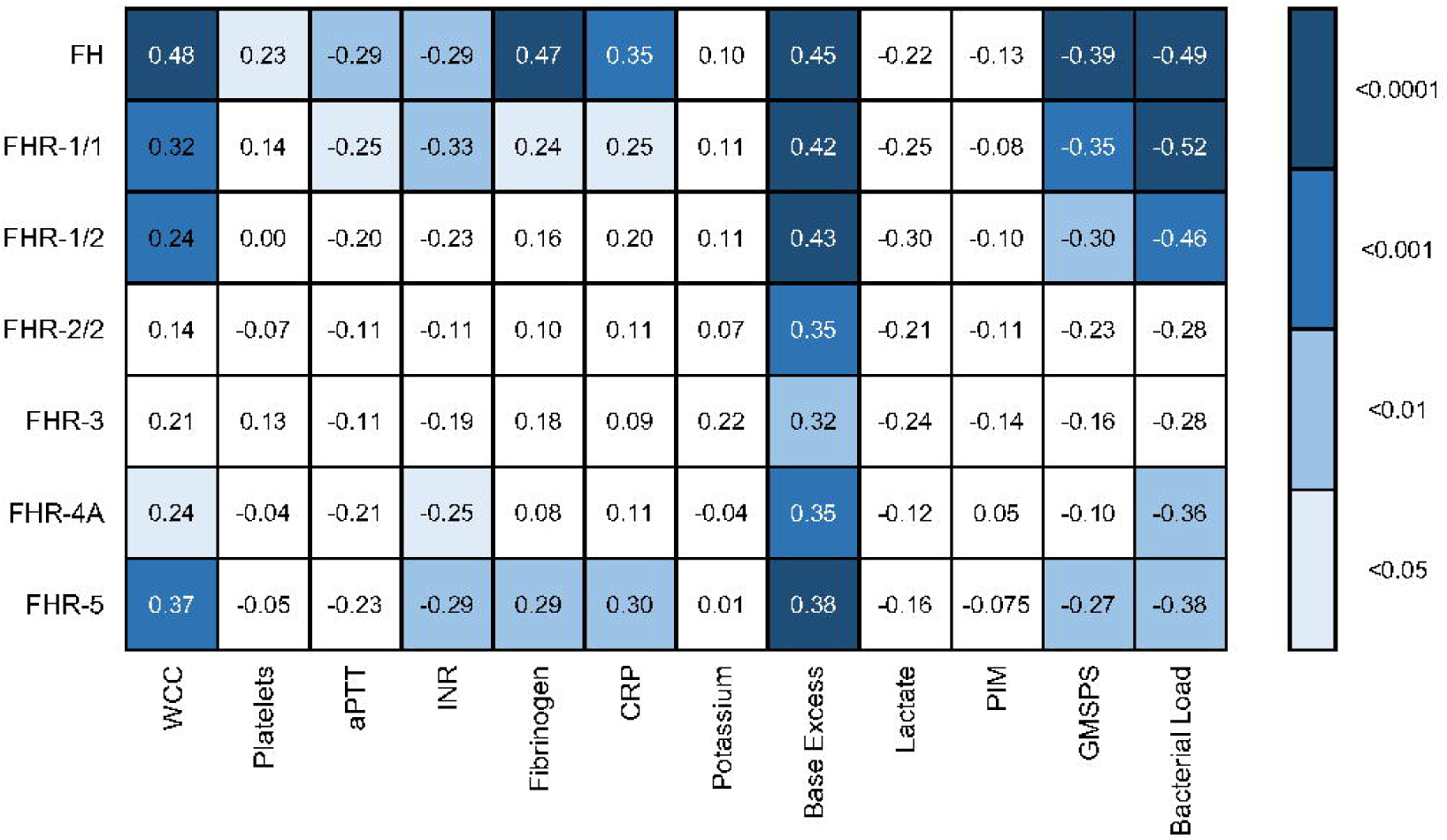
Associations of acute stage FH family protein levels with clinical and laboratory parameters. Pearson correlation coefficients (*r*), considering twelve severity markers and seven FH family proteins (including the different dimers). Correlations were assessed using Pearson’s measure of association, followed by the Benjamini-Hochberg procedure to control for the false discovery rate (FDR, set to 0.05). Blue shades indicate, from light to dark: *p* < 0.05; *p* < 0.01; *p* < 0.001; and *p* < 0.0001. WCC: white cell count; aPTT: activated partial thromboplastin time; INR: international normalized ratio; CRP: C-reactive protein; PIM: pediatric index of mortality; GMSPS: Glasgow Meningococcal Septicemia Prognostic Score.

**Figure 5.**
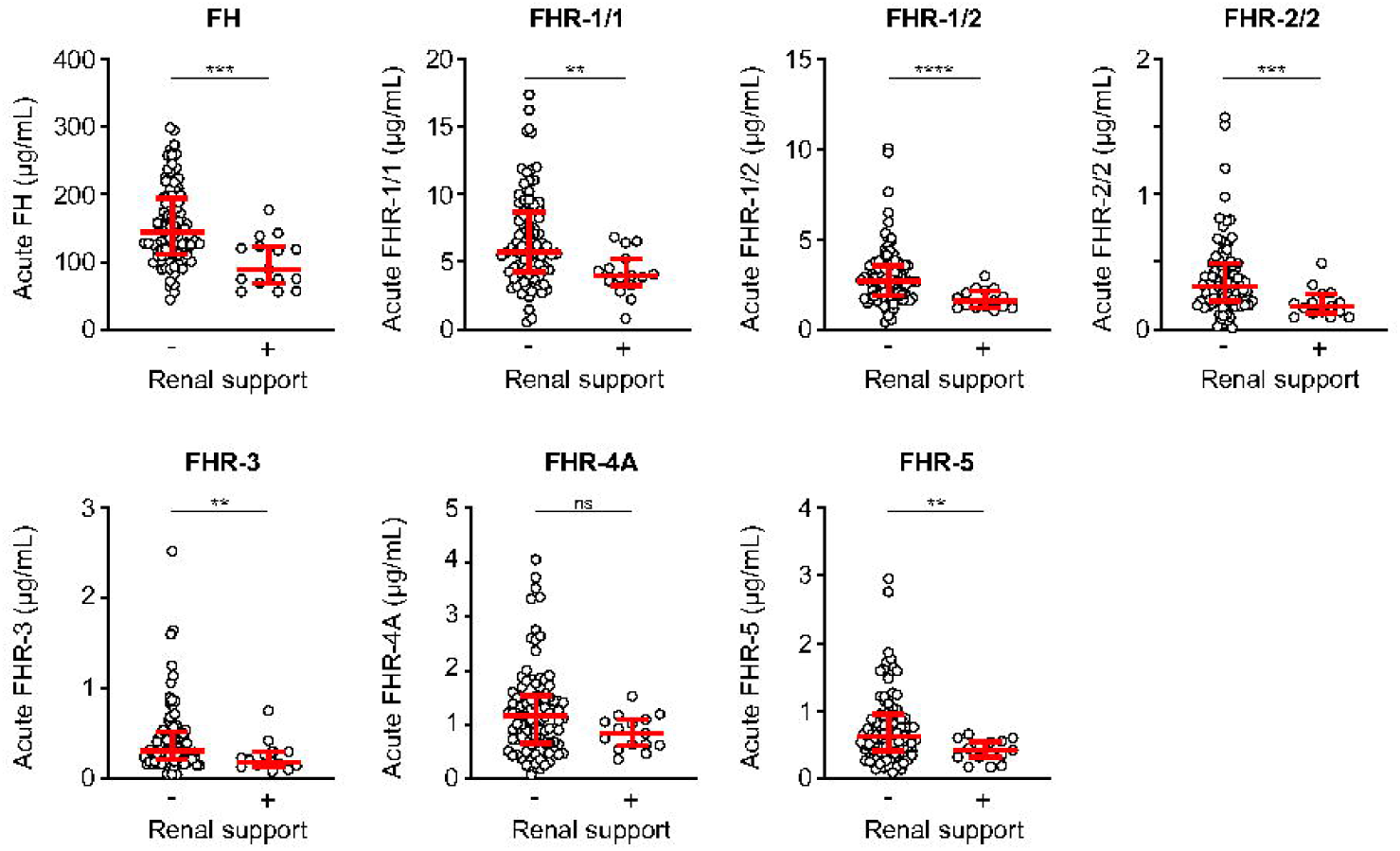
FH family proteins are low in patients who receive renal support. Serum levels of FH, FHR-1/1, FHR-1/2, FHR-2/2, FHR-3, FHR-4A and FHR-5 at the acute stage of patients who did (*n* = 15) or did not (*n* = 90) receive renal support. Both surviving and non-surviving patients are included here. Statistical significance was tested using a Mann-Whitney test. Lines depict median and IQR. ****: *p* < 0.0001; ***: *p* < 0.001; **: *p* < 0.01; *: *p* < 0.05; ns = not significant.

Acute stage levels FH, FHR-1/1, FHR-1/2, FHR-4A and FHR-5 correlated with bacterial load, which is a well-established marker of MD severity (**Figure 4**). We determined the *N. meningitidis* bacterial load in 62 patients of whom sufficient acute stage serum was available. The bacterial load ranged from 8.52*10^0^ to 1.04*10^9^ copies per mL serum (median = 5.68*10^4^). We did not observe a difference between serogroups B and C in bacterial load (*p* = 0.33, Mann-Whitney test), nor did we observe a difference in severity (by PIM score, *p* = 0.26, Mann-Whitney test). Except for FHR-5, we did not observe a significant association between *N. meningitidis* serogroups B or C and acute stage FH family protein levels (**Supplementary Figure 3**). Bacterial load correlated negatively with base excess (*r* = −0.42, *p* < 0.001) and positively with lactate (*r* = 0.68, *p* < 0.0001).

## DISCUSSION

The role of complement in MD is complex and can be regarded as a “double-edged sword”. While its activation is essential for the clearance of invading microbes, too much complement activation may contribute to immunopathology. Plasma levels of FH are subjected to a similarly delicate balance: High FH levels protect host tissues from complement-mediated damage, but this also increases survival of FH-binding pathogens such as *N. meningitidis*. In contrast, low FH levels decrease complement evasion of FH-binding pathogens but renders host tissue more vulnerable to complement-mediated damage. This delicate balance in FH levels is further complicated by the presence of the proposed FH antagonists, the FHRs. It is hypothesized that the competition between FH and the FHRs determines to what extent and at what rate alternative pathway activation takes place on surfaces [16]. With the recent insight that fHbp also binds FHR-3, it was suggested that the levels of FHR-3 determine if *N. meningitidis* successfully recruits FH and evades complement, and thereby would play a crucial role in developing MD [22]. Our previous research demonstrated that a systemic competition between FH and FHR-3 is unlikely to occur during steady-state, where the molar excess of FH is ∼130-fold compared with FHR-3 [8]. However, it was unknown up until now whether this would also hold true during MD.

Although FH and possibly FHR-3 play an important role in MD susceptibility, the dynamics of their plasma levels during the acute stage of MD were unknown. By measuring all FH family protein levels during MD in the first days after admission, we observed that the protein levels of both FH and the FHRs were markedly decreased during the acute stage of MD, whereas their recovery towards normal levels showed different kinetics. FH and FHR-5 levels slowly progressed to normal levels, suggesting a low synthesis rate of these proteins by the liver, or alternatively, that their consumption or loss from the circulation may be prolonged. FHR-1, FHR-2 and FHR-3 showed quicker recoveries within a few days after admission, while FHR-4A did not recover during the first four days. As the main production site of the FH protein family is the liver, the various observed recovery rates indicate that liver synthesis is not generally low during MD.

During the acute stage of MD, necrotic or damaged tissue, including the vasculature, will activate complement [27,28]. The reduction of FH and the FHRs in the circulation may therefore be due to the subsequent recruitment of these proteins to sites of complement activation. Alternatively, it can be hypothesized that the protein levels are low because of additional passive leakage into the tissues due to increased vascular permeability [29]. Both processes could account for the lower protein levels during the acute stage of meningococcal septicemia, which is a more systemic disease in comparison to meningococcal meningitis. In support of the increased vascular permeability as an underlying mechanism, the serum levels of IgG (of similar molecular weight as FH) were also decreased, suggesting a non-selective disappearance of proteins from the blood compartment. This is in line with clinical vascular leakage and edema formation known to occur during treatment at the early stage of the disease. If vascular leakage would be accounting completely for the initial loss of proteins, similar recovery rates are expected. However, the FH protein family showed different dynamics in subsequent days, suggesting that other processes are at play. We could not determine whether their different dynamics are reflecting different roles for each of the proteins during inflammatory responses or are due to different rates of synthesis.

Finally, FH levels could be low due to recruitment via fHbp on the meningococci. However, this was also observed for FHRs that are not bound by *N. meningitidis*. Moreover, the observed decreases of all FH family proteins correlated strongly with each other and were reduced to a similar extent as IgG. This suggests that the reduced plasma concentrations are due to general leakage from the circulation rather than consumption due to binding on meningococci.

FHR-3 is the only FHR bound by fHbp that can compete with FH in vitro and might affect *N. meningitidis* survival [22]. However, its acute stage protein levels did not correlate with bacterial load. The vastly different molar concentrations of FH and FHR-3, with FH circulating at a >100-fold excess compared to FHR-3, make it unlikely that substantial amounts of FHR-3 would be bound by meningococci, and that this would be reflected in plasma levels. The proposed role of FHR-3 as competitor of FH in meningococcal disease therefore is unlikely to play a role in the circulation.

Of the FH protein family, FH was found to correlate most with MD severity parameters. Lower FH levels during the acute stage correlated with the prediction score GMSPS as well as the blood variables which are known to be associated with MD severity, including WCC, platelets, base excess, INR and aPTT. The correlation of FH with coagulation markers such as fibrinogen, INR and aPTT is in line with a previous study that showed decreased levels of FH when sepsis was accompanied by disseminated intravascular coagulation [30], and corroborates an in vitro study that suggests a potential role for FH in coagulation [31]. The association of FH with severity also explains the observed association of FH and FHRs with bacterial load. While, as discussed above, this is unlikely to reflect consumption, bacterial load is a strong predictor of severity, and thus the associations with FH and FHRs are suggestive of FH and FHR clearance due to severe inflammation and the accompanying complement activation [32,33].

Low levels of most FH family proteins were associated with renal failure during hospitalization. While this may be part of the association of reduced FH and FHRs with disease severity, regulation of complement activation by FH is known to be involved in renal disease including hemolytic uremic syndrome and glomerulonephritis [16]. The association of FH and FHRs with impaired kidney function may not simply be due to disease severity, but rather may indicate a specific protective role of the FH family proteins in preventing renal injury.

The double activity of FH, protecting both host tissue and meningococci against complement-mediated damage, makes analysis of its role in MD challenging. Following what has previously been found in vitro for *N. meningitidis* [15], where small changes in FH levels greatly affect survival, we propose that a balanced concentration of FH in blood is key in MD. Low steady-state FH levels may reduce susceptibility towards MD, but if MD does occur, the low FH levels may be insufficient to protect the vasculature and kidneys from complement-mediated damage [34]. Indeed, it has previously been shown that inhibition of complement improves survival of mice suffering from bacterial meningitis [35]. Therapeutic interventions that substantially increase the levels of FH activity on host surfaces (and not on microbial surfaces) may reduce tissue injury, when administered as soon as patients have been treated with antibiotics to eliminate meningococci from the circulation. A potentiating antibody that increases the function of FH on host surfaces without enhancing binding to meningococci might therefore be worth investigating in acute stages of systemic inflammation [34].

## MATERIALS and METHODS

### Study cohort

Patients in this study (n = 106) were a subset of the cohort recruited at St. Mary’s Hospital, London (UK) between 1992 and 2003, the details of which have been reported previously [15,21,36–38]. All samples were obtained with informed consent of the parents or guardians of each patient according to the local ethics committee and the Declaration of Helsinki and were stored at −80 until use. Cases included in this study had microbiologically confirmed MD and had acute serum samples taken during hospitalization, and a serum sample taken after convalescence (in survivors, n = 91).

### Blood Markers of disease severity

White cell count (WCC), platelet count, activated partial thromboplastin time (aPTT), international normalized ratio (INR), base excess, and levels of fibrinogen, C-reactive protein (CRP), potassium, and lactate were all determined as part of routine diagnostics at St. Mary’s Hospital, London (UK). Glasgow Meningococcal Septicemia Prognostic Score (GMSPS) and Pediatric Index of Mortality (PIM) score were determined as previously described [39,40].

### Measurement FH and FHR proteins in serum

FH family proteins and total human IgG levels in serum were determined by in-house developed ELISAs as previously described [8,18,23,25]. In short, FH was measured by ELISA using an in-house generated, specific mouse monoclonal antibody (mAb) directed against CCP domains 16/17 of FH (clone anti-FH.16, Sanquin Research, Amsterdam, the Netherlands) as the capture mAb and polyclonal goat anti-human FH (Quidel, San Diego, CA, USA), which was HRP-conjugated in-house, as the detecting Ab. FHR-1/1 homodimers and FHR-1/2 heterodimers were captured using clone anti-FH.02 (Sanquin Research), and detected with biotinylated anti-FH.02 (Sanquin Research) or anti-FHR-2 (clone MAB5484, R&D Systems, Minneapolis, MN, USA), respectively. For the specific detection of FHR-3, an in-house developed mAb directed against FHR-3 and FHR-4A (clone anti-FHR-3.1, Sanquin Research) was used as the capture mAb. A biotinylated mAb directed against FHR-3 and FH (clone anti-FHR-3.4, Sanquin Research) was used as detecting mAb. To measure FHR-4A, a rat anti-mouse kappa mAb (RM-19, Sanquin Research) was coated on the plate before addition of the capture mAb (clone anti-FHR-4A.04, Sanquin Research), capturing specifically FHR-4A. The detecting mAb was a biotinylated polyclonal rabbit anti-human FHR-3 (Sanquin Research). FHR-5 was measured by using two specific mAbs: anti-FHR-5.1 as a capturing mAb and biotinylated anti-FHR-5.4 as detection mAb (both from Sanquin Research).

To determine whether fHbp affects the measurement of FH and FHRs by ELISA, *e*.*g*. by sterically hindering or otherwise affecting specific antibody binding to FH or any of the FHRs, the ELISAs were performed as described above, with purified fHbp (a kind gift of Prof. Christoph Tang, University of Oxford, UK) added during the sample step. A concentration of 100 µg/mL fHbp was used relative to 100% serum, representing an approximate 1-to-1 molar ratio with FH, using fHbp wildtype variants 1 and 3, as well as the fHbp variant 1 mutant that is unable to bind FH.

### *N. meningitidis* quantification

Three sets of primers/probes widely used in molecular diagnostics laboratories targeting three different conserved genes (*metA, sodC* and *tauE*) in *N. meningitidis* were used for bacterial load quantification by qPCR. Master mixes (LightCycler^®^ 480 II Master mix, 2X conc., Cat. nr. 04887301001, Roche, Basel, Switzerland) contained 2.5 µL primers (200 nM), 2.5 µL probe (100 nM) and 20 µL template DNA in a total reaction volume of 50 µL, following manufacturer’s recommendations. Initial denaturation of 95°C for 7 minutes was followed by 50 cycles of 95°C for 10 s, 60°C for 40 s, and 72°C for 1 s and a final cooling step at 40°C for 10 s. Quantification was estimated using *N. meningitidis* DNA Standards copies/µL (dilution series of 1.15×10^−6^ to 1.15×10^−3^) AMPLIRUN^®^ DNA Control (1.15×10^−7^ copies/µL. Amplification data were analyzed by instrument software (Roche).

### Statistical analysis

Data were analyzed using GraphPad Prism, version 8 (GraphPad software, La Jolla, CA, USA) and R version 3.5.0 [41].

## Data Availability

Data within the manuscript is available upon request from the corresponding author.

## ACKNOWLEDGMENTS

The authors would like to thank the patients participating in the study.

