## Supplemental Information for "Loss of Factor H family proteins associates with meningococcal disease severity"

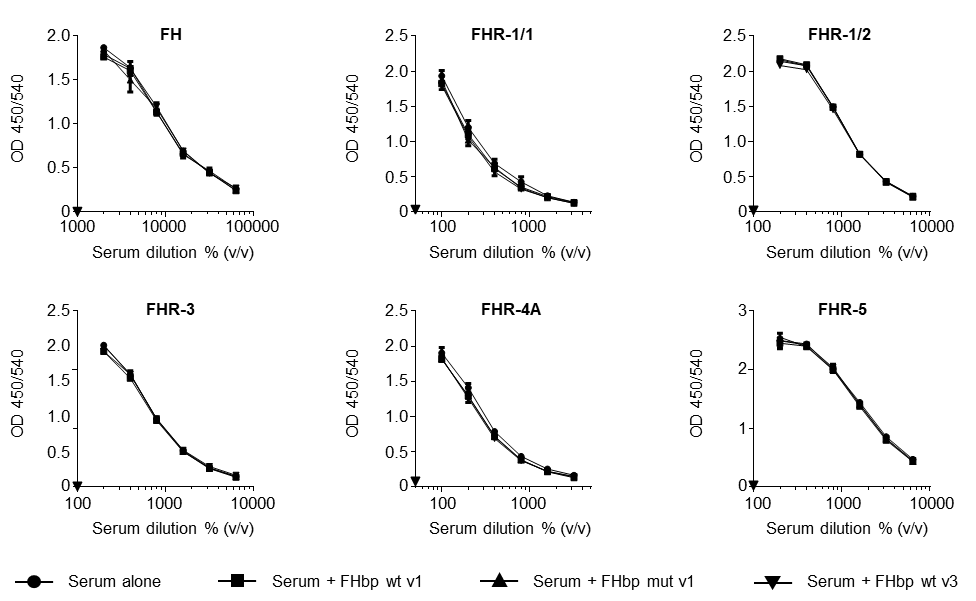

**Supplementary Figure 1 | no fHbp interference in the FH family protein ELISAs**. Serum level measurements of FH, FHR-1/1 homodimers, FHR-1/2 heterodimers, FHR-3, FHR-4A and FHR-5 were tested for interference by recombinant fHbp wildtype of variant 1 (wt v1) and variant 3 (wt v3). Recombinant fHbp that was unable to bind FH was used as control (mut v1).

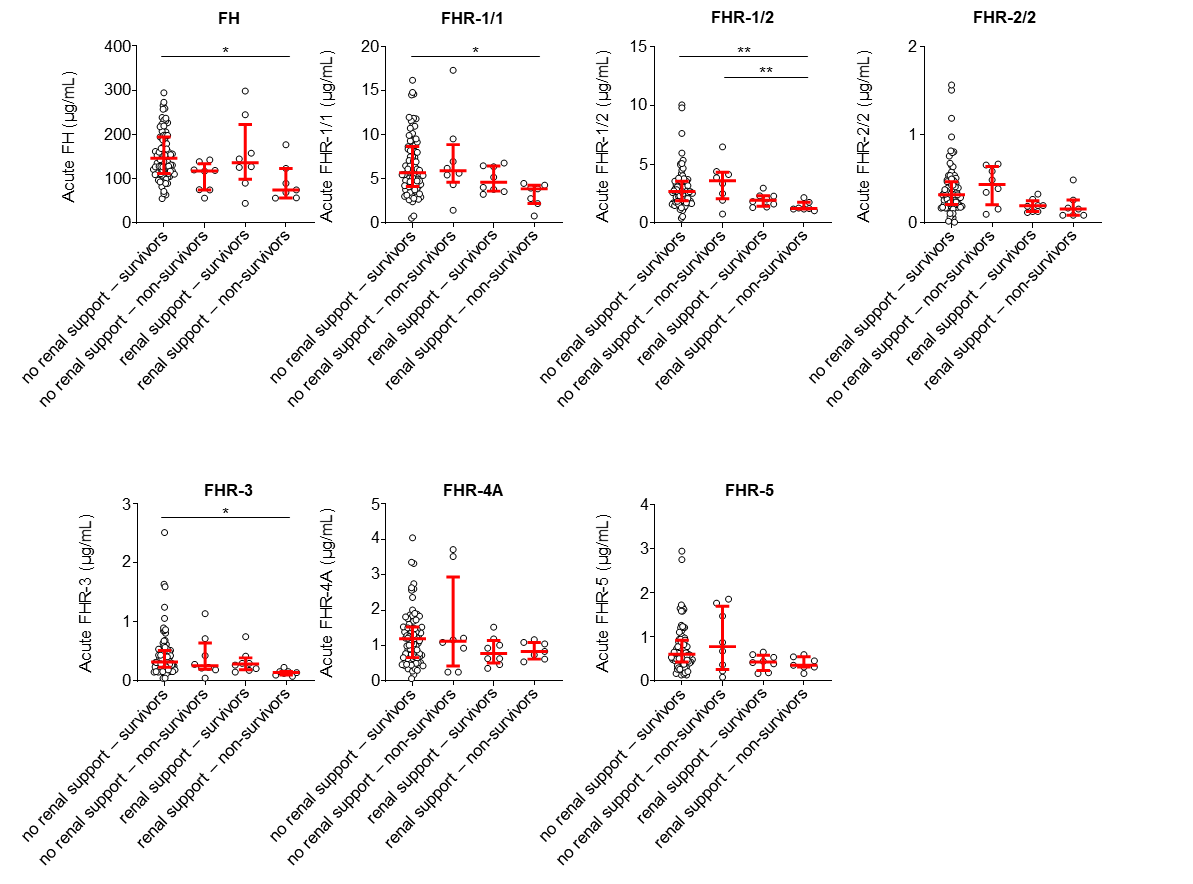

**Supplementary Figure 2 | The need for renal support in non-survivors.** Serum levels of FH, FHR-1/1, FHR-1/2, FHR-2/2, FHR-3, FHR-4A and FHR-5 at the acute stage of survivors who did (*n* = 8) or did not (*n* = 83) receive renal support and non-survivors who did (*n* = 7) or did not (n = 8) receive renal support. Statistical significance was tested using a Kruskal-Wallis test, followed by a Dunn’s multiple comparisons test. Lines depict median and IQR. **: *p* < 0.01; *: *p* < 0.05.

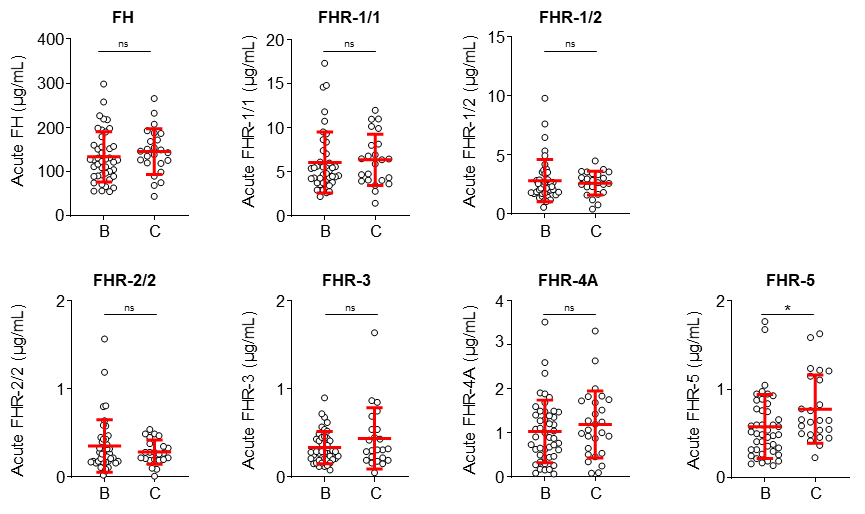

**Supplementary Figure 3 | FH family proteins per serogroup.** Serum levels of FH, FHR-1/1, FHR-1/2, FHR-2/2, FHR-3, FHR-4A and FHR-5 according to the meningococcal serogroup, being either serogroup B (*n* = 43) or serogroup C (*n* = 24). Statistical significance was tested using a Mann-Whitney test. Lines depict median and IQR. *: *p* < 0.05; ns = not significant.
